# Laboratory validation of an RNA/DNA hybrid tagmentation based mNGS workflow on SARS-CoV-2 and other respiratory RNA viruses detection

**DOI:** 10.1101/2020.05.12.20099754

**Authors:** Feili Wei, Yanhua Yu, Zhongjie Hu, Rui Wang, Xianghua Guo, Haiying Jin, Shan Guo, Yabo Ouyang, Ying Shi, Ronghua Jin, Dexi Chen

**Affiliations:** Beijing Institute of Hepatology, Beijing Youan Hospital, Capital Medical University,, Beijing, 100069, China; Beijing Precision Medicine and Transformation Engineering Technology Research Center of Hepatitis and Liver Cancer, Beijing, 100069, China; Clinical Laboratory Center, Beijing Youan Hospital, Capital Medical University, Beijing, 100069, China; Center for Infectious Diseases, Beijing Youan Hospital, Capital Medical University, Beijing, 100069, China; Department of Critical Care Medicine of Liver Disease, Beijing Youan Hospital, Capital Medical University, Beijing, 100069, China

**Author notes:** These authors contributed equally to this work. R. J and D. C contributed equally to this work. **Corresponding author:** D. Chen.

**Keywords:** RNA viruses, COVID-19, mNGS, Tn5 transposon, analytical validation

## Abstract

**Background:** Acute respiratory infection (ARI) caused by RNA viruses is still one of the main diseases all over the world such as SARS-CoV-2 and Influenza A virus. mNGS was a powerful tool for ethological diagnosis. But there were some challenges during mNGS implementation in clinical settings such as time-consuming manipulation and lack of comprehensive analytical validation.

**Methods:** We set up CATCH that was a mNGS method based on RNA and DNA hybrid tagmentation via Tn5 transposon. Seven respiratory RNA viruses and three subtypes of Influenza A virus had been used to test CATCH’s capabilities of detection and semiquantification. Analytical performance of SARS-CoV-2 and Influenza A virus had been determined with reference standards. We compared accuracy of CATCH with quantitative real time PCR by using clinical 98 samples from 64 COVID-19 patients.

**Results:** We minimized the library preparation process to 3 hours and handling time to 35 minutes. Duplicate filtered RPM of 7 respiratory viruses and 3 Influenza A virus subtypes were highly correlated with viral concentration (r=0.60, p<0.001, Spearman correlation test). LOD of SARS-CoV-2 was 39.2 copies/test and of Influenza A virus was 278.1 copies/mL Comparing with qR-TPCR, the overall accuracy of CATCH was 91.4%. Sensitivity was 84.5% and specificity was 100%. Meanwhile, there were significant difference of microbial profile in oropharyngeal swabs among critical, moderate patients and healthy controls (p<0.001, PERMANOVA test).

**Conclusion:** Although further optimization is needed before CATCH can be rolled out as a routine diagnostic test, we highlight the potential impact of it advancing molecular diagnostics for respiratory pathogens.

## Background

Respiratory tract infection, as a global burden on public health, has caused extensive morbidity and mortality worldwide in the past decades. It is 3rd leading causes of death among all ages by 2015^1^. In most cases of severe acute respiratory tract infections, RNA viruses are pathogenic agents^2-4^. For example, the novel coronavirus SARS-CoV-2, which is pathogen of COVID-19, has caused over 3 million people infected and thousands death from December 2019 until now^5,6^. Traditional nucleotide analysis methods for RNA viruses, such as quantitative reverse transcription PCR (qRT-PCR), are sensitive and specific, but only focused on pre-defined species or subtypes.

Metagenomic next-generation sequencing (mNGS), which is well studied on pathogen detection recently, offers a culture-free and nucleotide sequence independent method that eliminates the need to define the targets for diagnosis beforehand^7^. However, most of those protocols have time-consuming steps and complicated manipulation including RNA extractions, reverse transcription, second-strand complementary synthesis, preamplification/isothermal amplification, adapter ligation and PCR amplification. It not only takes a long time to wait for diagnostics results but also not easy for clinician to use in a large scale. Bacterial transposase Tn5 has been employed in next generation sequencing by cutting double-stranded DNA (dsDNA) and ligate the resulting DNA ends to specific adaptors^8-10^. It has been successfully used in mNGS for pathogen detection in low input specimens^11^. Furthermore, it also can skip second-strand complementary synthesis by direct tagmentation of RNA/DNA hybrids in scRNA-seq and SARS-CoV-2^12-14^. But the lack of method standardization and validation of these workflows encumbers the ability to assess the variability of results generated by different laboratories, leading to uncertainty in the results^7,11,15,16^. Hence, laboratory validation including low limit of detection (LOD), precision, stability, effects of interference and accuracy for RNA viruses in one single assay is essential for implementation of mNGS for routine pathogen detection in clinical diagnostic laboratories.

In this study, we optimized metatranscriptomic protocol based on RNA/DNA hybrid tagmentation for detecting respiratory RNA viruses. We call this method as **CATCH** (pathogeni**C** rn**A**/dna hybrid **T**agmentation te**CH**nology). We aimed to build up a rapid and accurate method for mNGS to be used as a broad diagnostic tool for viral respiratory diseases with the potential for pan-pathogen detection. Despite validation of laboratory performance, we also determined its sensitivity and accuracy compared to that of existing diagnostics method–qRT-PCR, using clinical samples from hospital patients during these COVID-19 pandemic. Further optimization is needed before CATCH can be rolled out as a routine diagnostic test, but we highlight the potential impact of it advancing molecular diagnostics for respiratory pathogens.

## Methods

### CATCH workflow for respiratory RNA viruses detection

RNA was extracted using QIAamp Viral RNA Mini Kit (Qiagen, Cat.No. 52904) following the manufacturer’s instructions. Internal RNA controls, consisting of an RNA phage (Escherichia coli bacteriophage MS2, ATCC15597-B1, Hecin Scientific, Inc), was added into all the samples before RNA extraction at a concentration of 1×10^3^ copies/mL, in approximately 10-fold of its LOD^11^. DNA was then removed through Denature buffer and DNase in Repli-g WTA kits (Qigen, Cat.No.150063). After removal of the DNA, RNA/DNA hybrids were synthesis by SuperScript^TM^ IV First-Strand Synthesis System (Thermo Fisher, Cat.No. 18091050). RNA/DNA hybrids tagmentation and indexing-PCR protocol are optimized according to Nextera XT DNA Library Prep Kit (Illumina, Cat.No. lFC-131-1096, Supplementary Protocol). For pooling, each library was quantified individually using the Qubit dsDNA HS Assay Kit (Thermo Fisher, Cat.No. Q32851), followed by combining equimolar concentrations of DNA libraries. The size distribution of the combined pools was determined using the High Sensitivity DNA kit (Agilent, Cat.No. 5067-5583) on an Agilent 4200 Bioanalyzer. Library pools were then sequenced on Illumina NextSeq 500 platform with SE (Single-End) 75 cycles sequencing strategy to generate a minimum of 10~20 million reads for each library.

### Bioinformatics workflow and data processing

We first used software fastp^17^ (v0.19.5) to filter low-quality reads and remove adapter with parameters: -q 20 -c -l 50), low complexity reads removal by Komplexity (-F, -k 8, -t 0.2, version: Nov-2019), host removal by bmtagger ftp://ftp.ncbi.nlm.nih.gov/pub/agarwala/bmtagger), and ribosomal reads removal by SortMeRNA^18^ (version:2.1b). Next, we applied Kraken2^19^ (version 2.0.8-beta, parameters: --threads 24 --confidence 0.2) to assign microbial taxonomic assignment against the large NT reference database (version: Nov-2019) combining with current SARS-CoV-2 reference genome (accession ID: NC_045512.2). Duplicate filtered RPM of each virus were calculated by the follow steps: 1) Extracting reads annotated as targeted viral species except for SARS-CoV-2. 2) Aligning all reads to targeted reference genome with tblastx^20^ (-max_target_seqs 1 -max_hsps 1 -evalue 1e-5, version:). Alignment with beyond 90% identity of reference sequence and 90% length of query reads are considered. 3) Reads with same start position were marked as duplicated reads and discarded. Rest of reads were used to calculate RPM (reads per millions) by normalizing sequencing data size. For SARS-CoV-2, reads belong to *Betacoronavirus* are extracted. Reference sequences of all *Betacoronavirus* were used for blastn^20^ alignment (-max_target_seqs 1 -max_hsps 1 -evalue 1e-5, version:). Best hits of SARS-CoV-2 have been kept for following analysis. The following steps were same with other viruses. For classification of Influenza A virus subtypes, TAEC^21^ had been used with default parameter and influenza viruses database from NCBI^22^. We defined positive detection of bacteria, fungus and viruses by the following threshold criteria: 1) more than 3 non over lapping reads from distinct genomic regions for viruses; 2) 10-fold higher duplicate filtered RPM than NTC. The rest of microbial taxon were considered as contamination and discarded.

### Study design of Respiratory RNA viral species and subspecies detection by CATCH

To study the performance of CATCH on detection of respiratory RNA viral species and subspecies, we designed a panel with two Reference Standards kit. The SARS-COV-2 Standard (Lot No. GBW(E)091090) was donated by the National Institute of Metrology, China and the 2nd National Reference Panel for Influenza A Viral Nucleic Acids Detection Kit (Lot No.370051-201801) was acquired from the National Institutes for Food and Drug Control, China. In brief, all of 7 Respiratory RNA viruses and 3 Influenza A virus subspecies standard reference were first tested its highest dilution fold (similar with LOD) except for SARS-CoV-2 and Influenza A virus (A/2009/H1N1) which had exact viral concentrations. We defined these concentrations as baseline for the other 6 RNA viruses and 2 Influenza A virus subspecies. For each virus or subspecies, we improved its concentration to 10-fold comparing with baseline. Each trial was repeated three times. For SARS-CoV-2 and Influenza A virus (A/2009/H1N1), we set 10^2^copies/test and 10^4^copies/mL as baseline. Each trial was repeated three times.

### Evaluation of analytical performance characteristics

**1) limit of detection:** SARS-COV-2 RNA standard was spiked into RNA extracted from oropharyngeal swabs VTM matrix of asymptomatic donors at concentrations ranging from 10 to 10^4^ copies/test. Influenza A virus (A/2009/H1N1) reference standard was spiked into oropharyngeal swabs VTM matrix at concentrations ranging from 10^2^ to 10^5^ copies/mL. LOD were calculated in R (version 3.5.1) using probit regression analysis^23^ following approved guidelines of clinical and laboratory standards institute with 3 to 20 replicates performed at each concentration. **2) Precision:** External PC and NTC samples were analyzed over 5 independent mNGS runs (inter-run reproducibility) and as 3 independent sets over 1 run (intra-run reproducibility) and evaluated for quality control metrics and viral detection using established thresholds. **3) Interference:** to evaluate the effect of human host background on mNGS assay performance, we spiked SARS-COV-2 and Influenza A virus (A/2009/H1N1) standards into low (1 × 10^4^ cells/mL), medium (1×10^5^ cells/mL), and high (1×10^6^ cells/mL) titers of human A549 cell. **4) Stability:** to assess stability, 3 replicates of external PC were placed in a refrigerator at 4°C for 0, 3 and 6 days. After these manipulations, mNGS libraries were generated from the samples and sequenced.

### Patients and clinical samples

Residual RNA samples from patients at Beijing Youan Hospital with laboratory confirmed COVID-2019 (diagnosed by qRT-PCR) and those with suspected COVID-2019 but testing negative. 64 patients were enrolled in this study according to the 7th guideline for the diagnosis and treatment of COVID-19 from the National Health Commission of the People’s Republic of China. Residual RNA samples (n=98) were stored at Youan prior to testing. mNGS test results were compared to qRT-PCR testing results. Sensitivity and specificity of the mNGS assay were calculated relative to prior qRT-PCR test. Moreover, oropharyngeal swabs from 15 healthy volunteers were collected and used as healthy controls.

### qRT-PCR assays

We performed qRT-PCR assay using the ABI ViiA7 (Applied Biosystems) instrument according to the CDC EUA-approved protocol. SARS-CoV-2 was detected with the following primers: For N gene, the forward 5’CACATTGGCACCCGCAATC3’, reverse 5’GAGGAACGAGAAGAGGCTTG3’ and the probe fam-5’ACTTCCTCAAGGAACAACATTGCCA3’-BHQ1. For ORF1ab gene, the forward 5’ GTGARATGGTCATGTGTGGCGG3’, reverse 5’ CARATGTTAAASACACTATTAGCATA3’ and the probe VIC-5’CAGGTGGAACCTCATCAGGAGATGC3’-BHQ1^24^. All oligos and probes were acquired from Sangon Shanghai.

### Statistical analysis

Mann Whitney U test or Kruskal Wallis rank sum test was used for continuous variables that do not follow a normal distribution. A comparison of microbiota was done by Perm-ANOVA test. Spearman method was used for correlation test between Ct values/viral concentration and duplicate filtered RPM.

### Ethics statement

This study was reviewed and approved by the Ethics Committee of Beijing Youan Hospital, Capital Medical University.

## Data Availability

The raw sequence data of samples in this study, after removal of human reads, will be deposited to Genome Warehouse in National Genomics Data Center and the sequence Read Archive database.

## Acknowledgement

We thank Dr. Bin Yang and Mr. Peizhi Li at Illumina China for providing a comprehensive suggestion on this project and assistance of high-throughput sequencing. The SARS-COV-2 Standard (Lot No. GBW(E)091090) was donated by the National Institute of Metrology, China.

## Author contribution

D. C, Z. H and R. J conceived the project; R.W, X. G, H. J and S. G conducted experiments; Y. O.Y, Y. S and Y. Y analyzed the data; F. W wrote the manuscript with the help from all other authors.

## Results

### CATCH workflow for clinical RNA viruses detection

To further adaptation of clinical uses, we minimized mNGS library preparation time and steps by upgrading reverse transcriptase from Superscript III to Superscript IV (Thermo Fisher), combing extension and adaptor-PCR in one step and reducing PCR extension time within each cycle. We minimized the total time on library preparation to 3 hours and on-hands time to 35 min (Supplementary Protocol, Figure 1A). For confirmation of this workflow on RNA viruses detection, we tested 7 respiratory RNA viruses which were common in CAP patients including SARS-CoV-2, Influenza A virus (A/2009/H1N1), Influenza B virus (B/Vitoria), respiratory syncytial virus, parainfluenza virus, mumps virus and rubella virus. All 7 RNA viruses were detected though this workflow (Figure 1B). Except for exploring targeted virus detected or not, we also tested performance of our workflow on semi-quantification of each RNA virus by increasing concentration of each virus to ten times comparing with baseline separately and calculating fold-change of duplicate filtered RPM (Methods). We assumed that we could observe a significant increase of viral signals in mNGS assay if our workflow can quantitatively measure concentration of RNA viruses in spike-in samples. The fold-change of duplicate filtered RPM are expected to be 10X. If then it indicates a strong capability for quantification of our workflow. We found that viral signals of these 7 RNA viruses were ranged from 0.18[0.04, 0.33] to 177.06[141.03, 213.09] as baseline and fold-change comparing with baseline were ranged from 5.76[2.96, 8.55] to 19.05[9.82, 28.29] (Table 1). As we thought, all of 7 RNA viruses had a significant fold change in its own trials and duplicate filtered viral RPM generated by CATCH are significantly correlated with viral conentraion (r=0.60, p=7.83e-16). However, parainfluenza virus and mumps rubulavirus showed a lower fold change than expectation (Mann Whitney U test, p=0.029 and p=0.022), while rubella virus showed significantly higher fold-change than expectation (Mann Whitney U test, p=0.049, Figure 1B). Further, to explore the performance of distinguishing and quantifying related sub-species, we tested three important subtype of Influenza A virus: pandemic A/2009/H1N1, A/H3N2 and A/H7N9. All these three influenza A viruses can be detected. However, except for H3N2, the other two subtypes had lower fold change of duplicate filtered RPM than our expectation (Mann Whitney U test, p=0.002, p=0.29 and p<0.001, Figure 1C, Table 1).

**Table 1.** Detection and quantification performance of 7 RNA viruses and 3 influenza A virus subtypes by CATCH

| Viral Name | Genome Size | GC% | 1X | 10X | Fold-change comparing with baseline (1X) | p-value |
| --- | --- | --- | --- | --- | --- | --- |
|  |  |  | duplicate filtered RPM | duplicate filtered RPM |  |  |
| SARS-CoV-2 | 3k | 38.0 | 20.15(3.02, 37.28) | 215.12(160.07, 270.16) | 11.78(8.76, 14.79) | 0.126 |
| Influenza A virus (A/2009(H1N1)) | 13k | 44.2 | 177.06(141.03, 213.09) | 2198.24(1471.33, 2925.15) | 12.41(8.31, 16.52) | 0.127 |
| Influenza B virus (B/Victoria) | 14k | 40.2 | 9.08(5.53, 12.62) | 126.99(71.07, 182.92) | 13.99(7.83,20.15) | 0.108 |
| Respiratory Syncytial Virus | 15k | 33.2 | 11.22(7.55, 14.88) | 213.73(110.10, 317.36) | 19.05(9.82, 28.29) | 0.051 |
| Parainfluenza virus | 15k | 37.2 | 12.37(5.84, 18.89) | 191.04(140.71, 241.37) | 15.45(11.38, 19.52) | 0.029 |
| Rubella virus | 9.7k | 69.6 | 17.76(12.37, 23.16) | 119.89(6.49, 17.49) | 6.75(3.65, 9.85) | 0.046 |
| Mumps rubulavirus | 15k | 42.5 | 20.05(8.38, 31.77) | 165.39(59.58, 171.92) | 5.76(2.96, 8.55) | 0.022 |
| Influenza A virus (A/2009(H1N1)) | 13k | 44.2 | 4.73(3.90, 5.55) | 16.49(10.46, 22.52) | 3.36(2.13, 4.59) | 0.002 |
| Influenza A virus (A/H3N2) | 13k | 45.0 | 5.25(2.95, 7.55) | 38.92(11.26, 66.58) | 8.09(2.34, 13.84) | 0.29 |
| Influenza A virus (A/H7N9) | 13k | 45.1 | 1.10(0.62, 1.57) | 5.36(3.08, 7.63) | 1.69(0.97, 2.40) | <0.001 |

**Figure 1.**
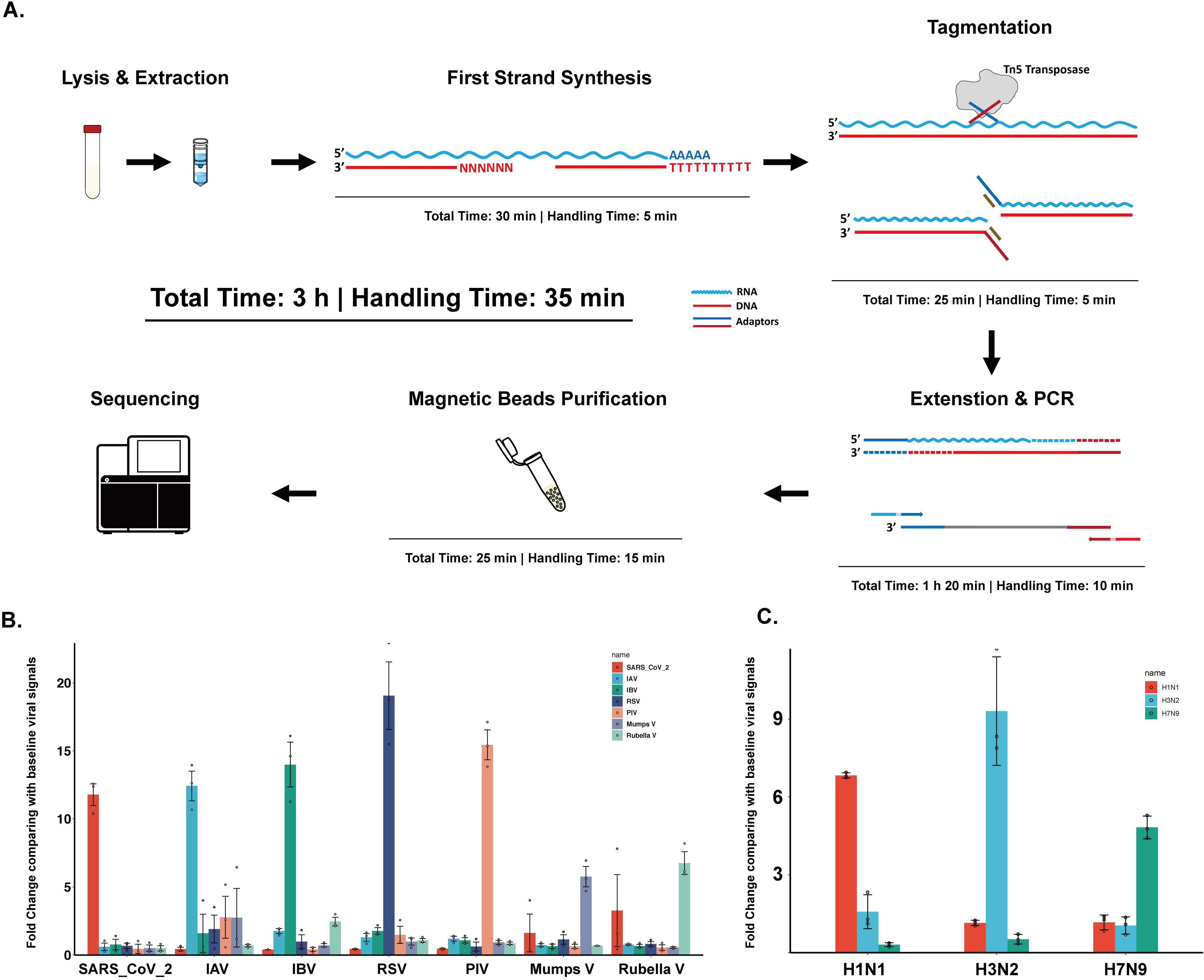
Schematic of RNA/DNA hybrid tagmentation workflow and its applications on RNA viruses detection. (A) Optimization of RNA/DNA hybrid tagmentation workflow. (B) Detection and semi-quantification of 7 typical respiratory RNA viruses. (C) Detection and semi-quantification of 3 typical Influenza A virus subtype. IAV: Influenza A virus, IBV: Influenza B virus, RSV: respiratory syncytial virus, PIV: parainfluenza virus, Mumps V: mumps virus and Rubella V: rubella virus.

### Analytical performance of Influenza A virus and SARS-CoV-2

To calculate 95% limits of detection (LOD), defined as the lowest concentration at which 95% of positive samples were detected, we evaluated SARS-CoV-2 and Influenza A virus separately at 5 different concentration levels, testing three to 20 replicates at each concentration. From 10^5^ copies/mL to 500 copies/mL library produced 2,163.00[1,093.23, 3,224.10] duplicate filtered RPM mapping to Influenza A virus. For SARS-CoV-2, we got 1,533.67[1,015.48, 2,049.86] to 11[6.03, 15.97] duplicate filtered RPM from 10^4^ copies/test to 50 copies/test (Figure S1A and B). Across the dilution series, duplicate-filtered RPM of both viruses were strongly associated with viral copies or titers (Spearman Correlation Test, p=1.41e10^-11^ and p=2.12e10^-11^). Using probit regression analysis, a 95% limit of detection is determined for SARS-CoV-2 and Influenza A virus (Table 2). Further, we evaluated the effects of interference from host cell on mNGS assay performance. Host cell at a level of more than 10^5^ cells /mL resulted in a significant reduction in number of IC and virial signals of Influenza A virus comparing with no additional host cell (Mann Whitney U test, p=0.06, p=0.04 and 0.03, Figure S1C). Moreover, inter- and intra-assay reproducibility and stability of held in 4°C for 0,3,6 days are also performed (Table 2). We did not find any significant difference among Days 0, 3 and 6 (Mann Whitney U test, p=0.23 and p=0.53 and 0.55, Figure S1D).

**Table 2.** Performance characteristics for CATCH on Influenza A virus and SARS-CoV-2

| Performance metrics | Method | Results |
| --- | --- | --- |
| Limits of detection | SARS-CoV-2 | 39.2 copies/test |
|  | Influenza A virus (A/H1N1/2009) | 278.1 copies/mL |
| Precision | Qualitative detection of both viruses over 5 consecutive runs | 100% concordance |
|  | Qualitative detection of both viruses over 3 replicates on the same runs | 100% concordance |
| Stability | Qualitative detection of Influenza A virus held at 4°C for 0, 3, 6 days | 100% concordance |
| Interference | Qualitative detection of Influenza A virus in Spiked host cell A549 (low, medium and high concentration) | Only in 10<sup>4</sup> cell/mL, all spiked ICs are detected above QC thresholds; All spiked Influenza A virus are detected above QC expect for 2 replicates at high concentration level |

### Retrospective validation on swabs specimens of COVID-19 patients using qRT-PCR and mNGS workflow

For evaluation of accuracy, a total of 98 RNA samples from 64 COVID-19 patients are tested using both mNGS assay and qRT-PCR. SARS-CoV-2 viral signals are present in 44 mNGS positive samples in 47 dual-gene positive qPCR results. At single-gene positive samples, 5 in 11 generated SARS-CoV-2 viral. No reads classified as SARS-CoV-2 are obtained from mNGS assay of 40 qPCR-negative samples. Besides, there were no other DNA or RNA viruses were considered as positive according our threshold in these samples. Overall, CATCH showed 84.5% sensitivity and 100% specificity compared to original clinical test results (Figure 3A). And if only consider dual-gene positive samples, the sensitivity will increase to 93.7%. On the other hand, we found viral duplicate filtered RPM of SARS-CoV-2 have strong correlation with Ct value of N gene (R^2^=0.754) rather than ORF1ab ((R^2^=0.326, Figure 3B and C). Although it inferred that CATCH might have 3’bias, we still found that CATCH had capability of RNA virus detection and semi-quantifying viral copies concentration in clinical samples.

### Microbial profile in COVID-19 patients and healthy control

To explore unique features of microbial profile in COVID-19 patients, we recruit another 15 healthy volunteers to collect their oropharyngeal swabs. In sum, we got 36,845.1[27,519.4, 46,170.7] microbial RPM in COVID-19 patients and 109,398.1[102,779.5, 116,016.8] microbial RPM in healthy control. Patients and healthy control can be clustered as three groups by top 50 microbial genus (Figure S2A). Jensen-Shannon divergence of all microbiome was used to calculate distance of three groups (Critical, Moderate and Healthy, Figure S2B). There was significant difference between COVID-19 patients (both critical and moderate) and healthy control (PERMANOVA test, p < 0.001). And between critical and moderate group, we also found a difference in microbiome of oropharyngeal swabs (PERMANOVA test, p = 0.013). To exclude batch effects on microbial profile data, we re-classified samples according to collection date and library preparation batches (Figure S2A). There were significant differences in clusters grouped by collection date and library preparation batches (PERMANOVA test, p < 0.001 and p < 0.001). But if only considering COVID-19 patients, we found less impact of collection date and library preparation on microbial profile (PERMANOVA test, p = 0.053 and p = 0.182). We found characteristic microbial taxon in healthy control such as *Streptococcus, Prevotella* and *Veilonella* that were common in human upper respiratory tracts (Figure S2A). For COVID-19 patients, we also found an enrichment of opportunistic pathogen such as *Candida* genus (Mann Whitney U test, p < 0.001, Figure S3A and C). For further confirmation of this result, we used culture methods to detect *Candida* genus via VITEK2^25^. There were 2 in 4 ICU patients that were detected *Candida* genus positive.

## Discussion

As of today, acute respiratory infection (ARI) caused by RNA viruses is still one of the main diseases all over the world. According to the study of GLIMP^26^, EPIC^4^ and CAP-China^27^, viral CAP diagnosis was 37.2% in Asia, 23% in the United States and 39.2% in China. To improve our capabilities of etiological diagnosis in infectious disease, mNGS assay has been applied and studied by many researchers and is gradually developed to be a powerful tool in the past few years^28^. However, there were some challenges had to be overcome during mNGS implementation in clinical settings such as laborious wet-lab manipulations and lack of analytical validation following with a standard guideline^7,28^. In this study, we set up a rapid, ease-to-use and sensitive clinical mNGS workflow--CATCH based on RNA/DNA hybrids tagmentation via Tn5 for respiratory RNA viruses detection. Besides, we provide a series data of its analytical performance of SARS-CoV-2 and Influenza A virus. To our knowledge, this is the first study that testing comprehensive performance of mNGS method based on RNA/DNA hybrids tagmentation for pathogen detection. We highly consider that this newfound characteristic feature of Tn5 will help mNGS implementation in clinical settings. Thus, solid data of accuracy and robustness of CATCH in pathogen detection are required and important for its clinical uses.

Of many mNGS workflow for pathogen detection, capability for quantification is still a challenge for many reasons^29-31^. Although CATCH cannot provide a well-defined linearity of quantification for RNA viral detection, we still found that viral signals detected by CATCH (duplicate filtered RPM) are strongly correlation with viral copies concentration or titers both in spiked trials and clinical samples (Figure 1A and B, Figure 2B and C). We considered that CATCH can be used as a semi-quantification tool for many clinical uses and comparing with other detection methods. On the other hands, CATCH can distinguish related species even subspecies. Identification of different subtype of Influenza A virus are important for clinical diagnosis^32^.

**Figure 2.**
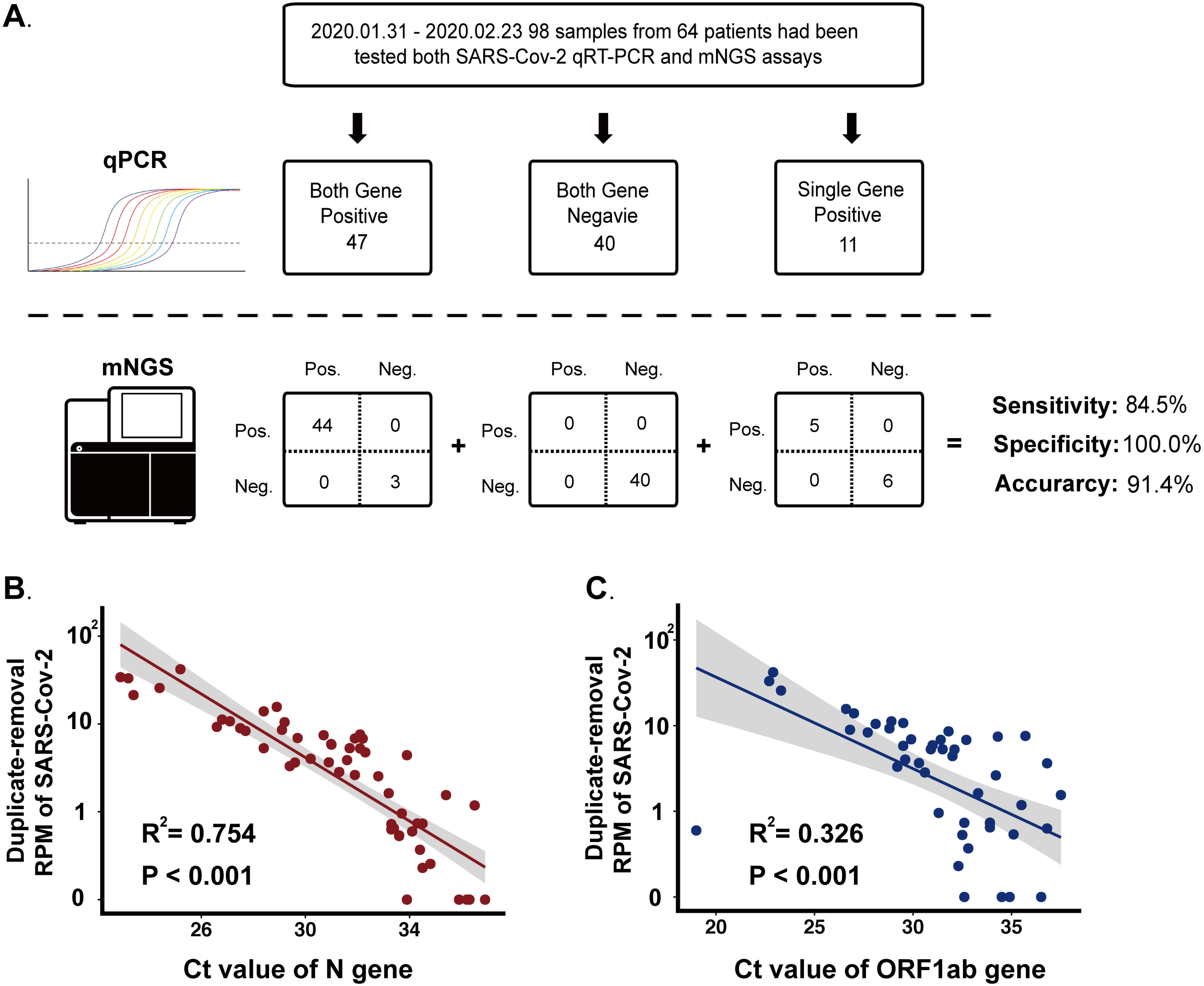
Accuracy of mNGS workflow comparing with qRT-PCR. (A) 2X2 contingency tables comparing the performance of mNGS workflow relative to qRT-PCR and clinical testing. The composite qRT-PCR standards used are both gene are detected (left), both gene negative (middle) and single gene detected (right). (B) Comparation of qRT-PCR ORF1ab gene Ct value and mNGS viral signals. (C) Comparation of qRT-PCR N gene Ct value and mNGS viral signals.

In analytical performance test with reference standard, we first found that limits of detection of CATCH were 39.2 copies/test for SARS-CoV-2 and 278.1 copies/mL for Influenza A virus. The sensitivity of Influenza A virus was at the same level of magnitude described in previous mNGS study either at Illumina platform^11^ or Nanopore platform^16^. While for SARS-CoV-2, we suggest that small genome size (three partial genomic regions, 3kb) might be the reason that CATCH cannot achieve a sensitive result the same as qRT-PCR^11,15^. Second, our data showed that high host background was a fairly common limitation of mNGS. It inferred that negative findings of unbiased mNGS methods might be less useful for excluding infection in high host background samples^11,16^. We, third, explore accuracy of CATCH by clinical oropharyngeal swabs during COVID-19 pandemic. CATCH showed a well compatibility with concentration of input. Although there were 65.00% and 38.46% samples in qRT-PCR negative and positive samples cannot be measured a definite concentration (Figure S3), we still have a 100% success in library preparation (Data not shown). The overall accuracy of CATCH for SARS-CoV-2 detection relative to conventical qRT-PCR was 91.4%, with 84.5% sensitivity and 100% specificity. In dual-gene qRT-PCR positive samples, sensitivity increased to 93.7%. We suggest that integrity of viral RNA might impair pathogen detective efficiency of CATCH. Moreover, as we knew, CATCH or other RNA/DNA hybrid tagmentation method without optimization will have obvious 3’bias^13^. This feature might also decrease possibility to capture viral fragments in clinical samples. For further implementation, CATCH needs optimization for overcome this limitation. Last, we found that CATCH can detect enrichment of opportunistic pathogen in oropharyngeal swabs of COVID-19 patients comparing with healthy control. It indicated that we can expand application of CATCH in fungal or bacterial detection in the near future. We also found an interesting phenomenon that microbiome in oropharyngeal swabs among critical, moderate patients and healthy people can be grouped in three clusters. Although we excluded batch effects to some extent such as collection date and library preparation batch, we still need more clinical data to confirm that there were significant difference in microbiome of oropharyngeal swabs between COVID-19 patients and healthy people for the reason that microbial profile might be affect by many artificial factors^33^. But it inferred that CATCH have a strong potential on finding microbial biomarker for disease evaluation and estimation^34-36^.

In summary, technological advancements in library preparation methods, sequence generation and computational bioinformatics are enabling quicker and more comprehensive metagenomic analyses at cost-effectiveness level. We hope CATCH will be a routine implementation of clinical mNGS in patient care settings.

---

Figure S1. Analytical characteristics of SARS-CoV-2 Influenza A virus. (A) LOD of SARS-CoV-2. (B) LOD of Influenza A virus. (C) Interference test with A549 host cell on Influenza A virus and Internal control. (D) Stability test on Influenza A virus and Internal control.

Figure S2. Microbial profile in oropharyngeal swabs of critical, moderate patients and healthy volunteers. (A) Heatmap of Top 50 microbial genus of critical, moderate patients and healthy volunteers. Red color represents high relative abundance and black color represents low relative abundance. (B) PCoA of microbiome in oropharyngeal swabs. (C) Relative abundance of *Candida genus* in different group.

Figure S3. RNA concentration of clinical oropharyngeal swabs specimens after DNase treatment. (A) Distribution of RNA concentration of throat swabs. Points lower than LOD of Qubit are generated by random function. (B) Proportion of samples with RNA concentration lower than LOD of Qubit.

